# Novel *FARS2* variants in patients with early onset encephalopathy with or without epilepsy associated with long survival

**DOI:** 10.1101/2019.12.30.19016139

**Authors:** Giulia Barcia, Marlène Rio, Zahra Assouline, Coralie Zangarelli, Charles-Joris Roux, Pascale de Lonlay, Julie Steffann, Isabelle Desguerre, Arnold Munnich, Jean-Paul Bonnefont, Nathalie Boddaert, Agnès Rötig, Metodi D. Metodiev, Benedetta Ruzzenente

## Abstract

Mitochondrial translation is essential for the biogenesis of the mitochondrial oxidative phosphorylation system (OXPHOS) that synthesizes the bulk of ATP for the cell. Hypomorphic and loss-of-function variants in either mitochondrial DNA or in nuclear genes that encode mitochondrial translation factors can result in impaired OXPHOS biogenesis and mitochondrial diseases with variable clinical presentations.

Compound heterozygous or homozygous missense and frameshift variants in the *FARS2* gene, that encodes the mitochondrial phenylalanyl-tRNA synthetase, are commonly linked to either early-onset epileptic mitochondrial encephalopathy or spastic paraplegia. Here, we expand the genetic spectrum of *FARS2*-linked disease with three patients carrying novel compound heterozygous variants in the *FARS2* gene and presenting with spastic tetraparesis, axial hypotonia and myoclonic epilepsy in two cases.

## Introduction

Mitochondria contain their own translation system complete with a mitochondria-specific ribosome (mitoribosome) and translation factors that function like their cytosolic counterparts. Intramitochondrial translation of mtDNA-encoded proteins is essential for the biogenesis of four of the five multiprotein complexes that form the ATP-producing oxidative phosphorylation system (OXPHOS). Inhibition of mitochondrial translation results in isolated or multiple OXPHOS deficiencies and mitochondrial disease.

*FARS2* (OMIM# 611592) encodes the mitochondrial phenylalanyl-tRNA synthetase (mtPheRS), which charges mt-tRNA^Phe^with phenylalanine for translation. MtPheRS contains four major domains: an N-terminal domain, a catalytic domain, a linker region (residues 290–322) and an anticodon-binding domain (ABD). Their complex interactions and conformational changes enable mtPheRS to function as a monomer during aminoacylation (1, 2). To date, 32 *FARS2* variants have been linked to two major clinical presentations: a) early-onset epileptic mitochondrial encephalopathy in about two-thirds of the cases (3-12); and b) spastic paraplegia (13-15). While the former group has a typically poorer prognosis, the latter is associated with less severe disease and prolonged survival. MRI studies of patients with early-onset disease often reveal global brain atrophy, cerebellar and basal ganglia abnormalities. In the second group, spastic paraplegia is associated with weakness of the lower extremities, spasticity and difficulties with walking. Intellectual disability/developmental delay may be observed in both groups. Here, we report the identification of 4 novel missense or frameshift variants in the *FARS2* gene in three unrelated patients (Pts) presenting with spastic tetraparesis, developmental delay and myoclonic epilepsy in two cases.

## Subjects and methods

Informed consent was obtained for all patients in accordance with the Declaration of Helsinki protocols and approved by local institutional review boards in Paris.

Detailed clinical presentations of the patients are shown in Table 1. Briefly, all three patients (Pts 1, 2 and 3) were born to non-consanguineous parents (Figure 1A) and presented with disease within the first weeks of life. They all had developmental delay and axial hypotonia. Pts 1 and 2 developed spastic quadriplegia. Pts 2 and 3 developed pharmaco-resistant epileptic seizures, mainly myoclonic and associated with a marked photosensitivity in Pt 2; brain MRI showed variably severe ventriculomegaly (Pts 1, 2 and 3) and brain-stem and basal ganglia involvement (Pts 2 and 3) (Figure 1B).

**Table 1.** Clinical characterization of Pts1, 2 and 3.

| Patient, sex, origin, age at the follow-up | FARS2 variants | Clinical summary at onset and during follow-up | Brain MRI | Biochemistry |
| --- | --- | --- | --- | --- |
| **Pt1**,<br>Female,<br>French and Chinese origin<br>8 years | Compound heterozygous<br>c.[1256G>A];<br>[1269_1276dup]<br>p.(Arg419His);<br>(Ser426*) | Regular pregnancy. BW: 2,471g (<5th percentile), BL: 44.5 cm (<5th percentile); BHC: 32.5cm (<5th percentile); Apgar score 7-9.<br>Presentation < 1 month with axial hypotonia and developmental delay; Head control was acquired at 10 months and she could sit at 2 years.<br>No signs of liver failure. No epileptic seizures.<br>EEG recording (repeated during the follow-up): normal background activity and physiological organization during sleep and wakefulness.<br>Electromyography and nerve conduction were normal.<br>At 8 years: spastic tetraparesis, dystonic movements, and axial hypotonia. Unable to walk unassisted. Dysarthric language, able to write using a keyboard.<br>No psychomotor regression episodes, but stagnation periods concomitant with febrile episodes during the first 3 years of life. | Normal at 1 and 2.2 years<br>Mild ventriculomegaly, no enlargement of the subarachnoid spaces (3.5 years) | Transient increase in PL: 1.4-2.9 mmol/l (normal range 0.7-1.8)<br>PP: 0.12-0.15 mmol/l (normal range 0.08-0.17)<br>Lactate/pyruvate ratio: 16-19 (normal range 6-14)<br>OXPHOS: ↓CI in muscle<br>↓CI+IV in fibroblasts |
| **Pt2**,<br>Female,<br>French origin,<br>16 years | Compound heterozygous<br>c.[989G>A];<br>[1113G>T]<br>p.(Arg330His);<br>(Leu371Phe) | Regular pregnancy. Normal growth parameters at birth; Apgar score 10/10. 6 weeks: neurological distress, global hypotonia.<br>No signs of liver failure.<br>Severe psychomotor delay. No head control. No visual contact. Can react to auditive stimuli.<br>From 19 months, started to present myoclonic focal and generalized seizures that were resistant to antiepileptic treatment composed by phenytoin, vigabatrin, topiramate, lamotrigine, and clonazepam in variable combinations.<br>EEG showed spikes and poly-spikes discharges predominating on occipital regions, activated by intermittent photic stimulation (IPS).<br>At 16 years: spastic tetraparesis and severe muscular atrophy predominating on inferior limbs. Mild scoliosis. | Marked ventriculomegaly, enlargement of the subarachnoid spaces due to white matter loss, especially in the sylvian fissures, abnormal T2 hyperintensities in the lentiform nuclei and dorsal brainstem (4 months, 15 years), cerebellar atrophy. Proton MR spectrum from the basal ganglia shows elevated lactates (15 years). | PL 2.5-4.8 mmol/l (<2.20 mmol/l)<br>PP: 0.21 mmol/l (normal range 0.08-0.17)<br>Lactate/pyruvate ratio: 13.7-16.6 (normal range 6-14)<br>CSF -L: 9.3 mmol/l (normal range: 1.4-2.2)<br>OXPHOS: ↓CIV in muscle and fibroblasts |
| **Pt3**,<br>Male,<br>French origin,<br>5 years | Compound heterozygous<br>c.[1256G>A];<br>[251A>C]<br>p.(Arg419His);<br>(His84Pro) | Regular pregnancy.<br>Normal growth parameters at birth; Apgar score: 10/10.<br>Global hypotonia since birth.<br>No signs of liver failure.<br>Severe psychomotor delay. Partial head control at 10 months.<br>Myoclonic generalized and focal seizures since the first year of life that were partially controlled by levetiracetam, clonazepam and piracetam.<br>EEG showed spikes and poly-spikes discharges predominating on occipital regions.<br>At 5 years: global hypotonia and lumbar mild scoliosis. He cannot sit unaided, has good contact, but no verbal language.<br>He has nasogastric feeding. | Moderate ventriculomegaly and enlargement of the subarachnoid spaces.<br>Dentate nuclei, brainstem, and pallidal T2 hyperintensity at the age of 1 and 3 years.<br>Normal spectroscopy. | PL: 2.6-4.8mmol/l (normal range 0.7-1.8)<br>PP: 7.8 mmol/l (normal range 0.08-0.17)<br>CSF-L: 5.8mmol/l (normal range: 1.4-2.2)<br>Normal liver enzymes<br>OXPHOS: ↓CIV in fibroblasts |

Accession numbers: FARS2 cDNA: NM\_001318872.1; mtPheRS: NP\_001305801.1, The normal ranges for the different biochemical indicators are indicated in the brackets.
List of abbreviation: BW, birth weigh; BL, birth length; BHC, birth head circumference; CSF-L, lactate in cerebral-spinal fluid; PL, plasma lactate; PP, plasma pyruvate.

**Figure 1.**
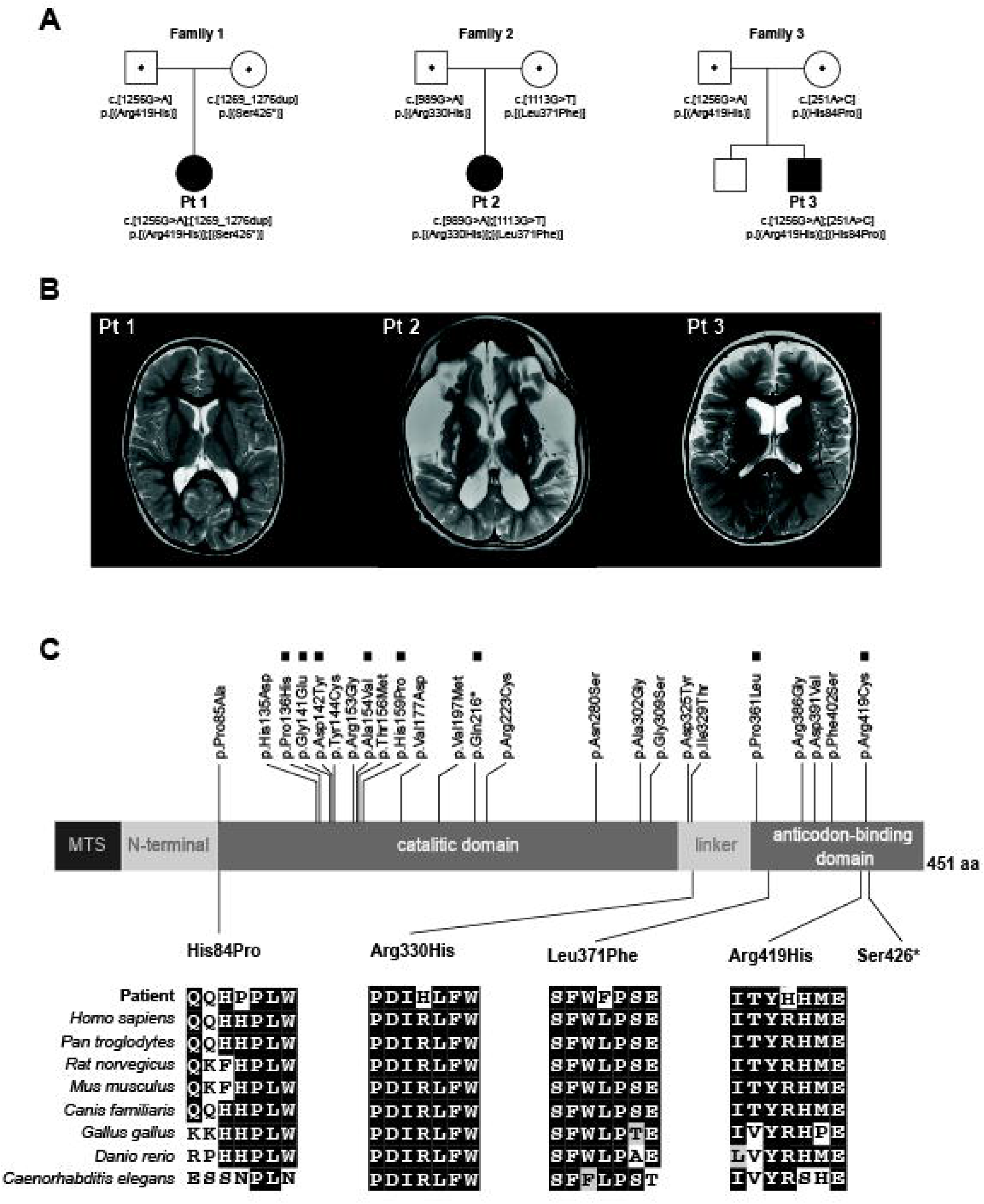
Family pedigrees, neuroimaging findings in patients carrying *FARS2* variants and evolutionary conservation of the amino acids affected by these variants. (A) Segregation of the newly identified *FARS2* variants into the pedigrees of the three reported families. (B) Axial T2-weighted images showing mild ventriculomegaly, no enlargement of the subarachnoid spaces in Pt 1; marked ventriculomegaly, enlargement of the subarachnoid spaces due to white matter loss, especially in the sylvian fissures, abnormal T2 hyperintensities in the lentiform nuclei and dorsal brainstem in Pt 2; moderate ventriculomegaly and enlargement of the subarachnoid spaces in Pt 3. (C) Linear map of the mtPheRS protein indicating its domain organization and position of the identified variants. Previously reported variants are shown above the mtPheRS map whereas variants reported here are below. FARS2 variants found in patients with hereditary spastic paraplegia are indicated with filed square. Evolutionary conservation of the amino acids affected by the newly described missense variants is shown in a multisequence alignment of mtPheRS proteins from different species.

Causative variants were identified using a next generation sequencing panel targeting 382 genes involved in mitochondrial diseases (Pts1 and 3) or whole exome sequencing (Pt 2). The variants were assessed *in silico* and confirmed by Sanger sequencing as in (16). Segregation analysis confirmed the presence of the variants in the parents.

Skin fibroblast cultures from controls and patients, protein extraction and immunoblotting were carried out as in (17). MtPheRS antibody was purchased from Proteintech (Manchester, UK, 16436-1-AP). The remaining antisera used here were described previously (17). *In vitro* pulse-labeling of mtDNA-encoded proteins was carried out in cultured patient fibroblasts as described in (17).

## Results

All three patients carried compound heterozygous variants including four missense variants NM_001318872.1:c.1256G>A p.(Arg419His), NM_001318872.1:c.989G>A p.(Arg330His), NM_001318872.1:c.1113G>T p.(Leu371Phe) and NM_001318872.1:c.251A>C p.(His84Pro) all affecting evolutionary conserved amino acids and predictably pathogenic, and one duplication NM_001318872.1:c.1269_1276dup p.(Ser426*) (Figure 1A and C, and Supplemental Table S1). None of these variants, but c.1256G>A, were previously reported in the literature. All newly identified variants were submitted to ClinVar (Submission ID).

Immunoblotting of whole-cell extracts revealed a decreased abundance of mutant mtPheRS in all three patients’ fibroblasts suggesting a structurally destabilizing role of the identified variants (Figure 2A). Examination of the abundance of mtDNA and nuclear-encoded OXPHOS proteins revealed a pattern of changes in their abundance consistent with impaired mitochondrial translation—decreased abundance of the mtDNA-encoded COXII protein and some, but not all, nuclear-encoded OXPHOS proteins (17), revealing multiple OXPHOS deficiency where CI and CIV are affected (Figure 2B). Interestingly, unlike COXII, the abundance of the mtDNA-encoded ATP8 was unchanged, probably because it contains a single phenylalanine whereas COXII contains ten (Figure 2B). Consistent with the immunoblot analysis above and the essential role in mtPheRS in mitochondrial translation, *in vitro* pulse labelling of mtDNA-encoded proteins revealed a general decrease in mitochondrial translation in fibroblasts from all three patients (Figure 2C). Cumulatively, these data indicate that the newly identified *FARS2* variants result in a destabilized mtPheRS protein and in an inhibition of mitochondrial translation.

**Figure 2.**
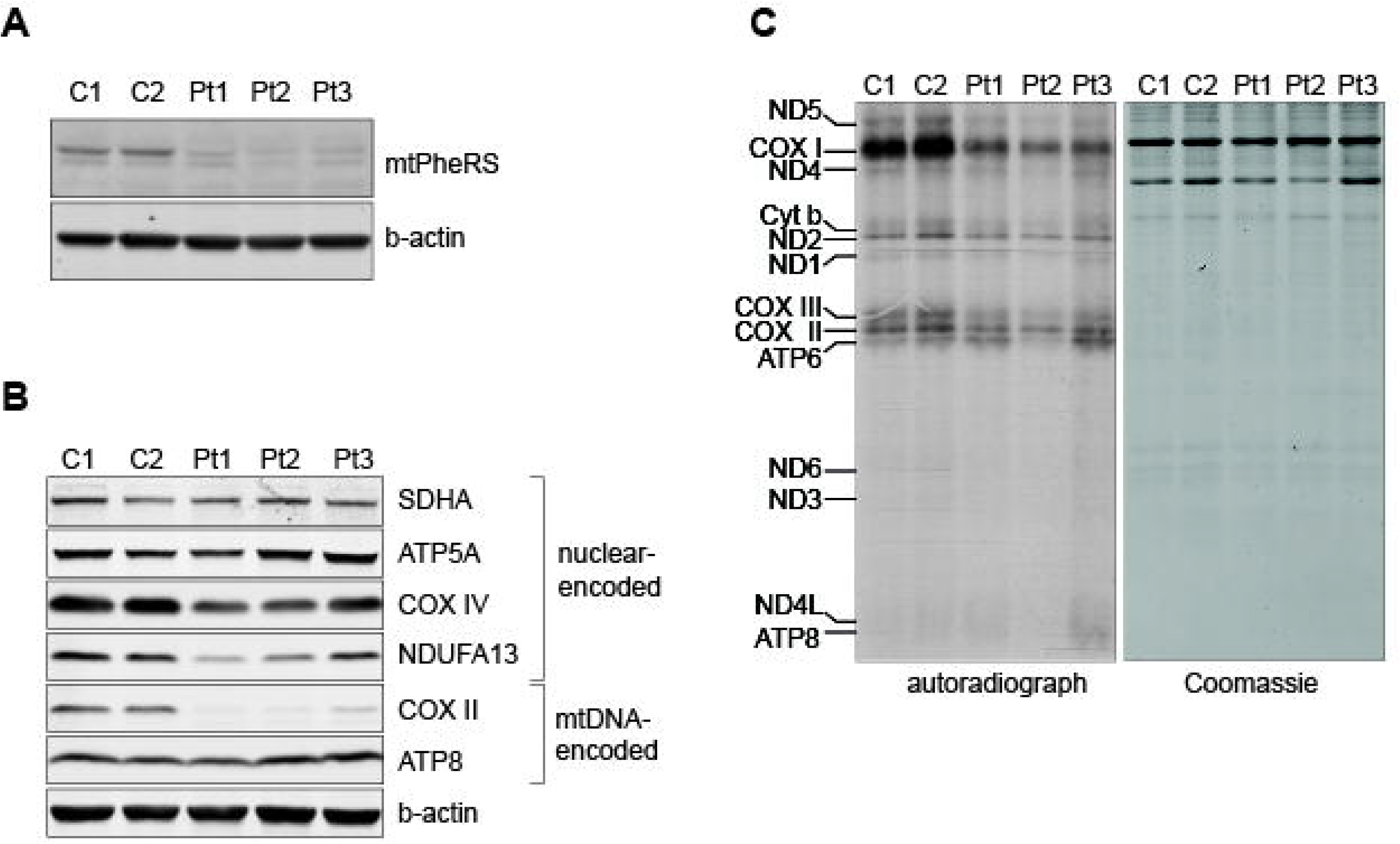
Decreased abundance of mtPheRS protein and an impaired mitochondrial translation in patient fibroblasts expressing different *FARS2* variants. (A) Immunoblot analysis of whole-cell extracts from control (C1 and C2) and patient (Pt 1, Pt 2, Pt 3) fibroblasts showing decreased abundance of mtPheRS. β-actin was used as a loading control. (B) Immunoblot analysis of whole-cell extracts from control (C1 and C2) and patient (Pt 1, Pt 2, Pt 3) fibroblasts showing decreased abundance of mtDNA-encoded COXII (OXPHOS complex IV) and nuclear-encoded NDUFA13 (OXPHOS complex I) and COXIV (OXPHOS complex IV). Abundance of SDHA (OXPHOS complex II), ATP5A (OXPHOS complex V) and the mtDNAencoded subunit ATP8 was unchanged. β-actin was used as a loading control. (C) Pulse-labeling of mtDNA-encoded proteins in control (C1 and C2) and patient (Pt 1, Pt 2, Pt 3) fibroblasts reveals an inhibition of mitochondrial translation in patient fibroblasts. Coomassie staining was used to assess loading. A representative of two independent experiments is shown.

## Discussion

Our report expands the genetic spectrum of *FARS2*-linked pathologies with four novel *FARS2* missense or duplication variants whose causal role was supported by bioinformatics and biochemical analyses.

The clinical presentations of *FARS2*-linked disease fall into two categories: a) early-onset severe encephalopathy with or without epilepsy or b) spastic paraplegia. The epileptic seizures observed in *FARS2* patients can have variable features in the different patients including neonatal multifocal seizures, infantile spasms or myoclonic epilepsy with an onset later in infancy. Patients with early-onset *FARS2*-linked encephalopathy also present with developmental delay and may have a wide range of brain abnormalities. Most of the previously reported *FARS2* patients with an early-onset encephalopathy died before two years of age (3-10, 12) whereas two patients with juvenile-onset epilepsy died soon after it became refractory (11). The clinical presentation of the three patients reported here was typical of *FARS2* disease with early-onset encephalopathy with or without epilepsy, included developmental delay and was associated with a prolonged survival. Pts 2 and 3 developed myoclonic pharmaco-resistant seizures whereas no seizures were reported for Pt 1. In contrast to most other patients with early-onset epilepsy, Pts 2 and 3 were alive at the age of 16 and 5 years, respectively. Similar finding has been reported for one other *FARS2* patient who presented with generalized epilepsy at age of 19 months and was alive at 17 years of age (9) suggesting that early-onset epilepsy can be associated with prolonged survival.

Even though hypomorphic or loss-of-function variants in different mitochondrial tRNA synthetases consistently result in neurological phenotypes, these are generally not specific to individual enzymes including *FARS2*—early-onset epilepsy and hereditary spastic paraplegia are genetically heterogeneous and not necessarily linked to tRNA aminoacylation. Moreover, there is no apparent correlation between the location of different *FARS2* variants along the mtPheRS polypeptide and the type, onset and severity of disease as, for example, variants affecting the catalytic domain are just as likely to cause an epileptic phenotype as variants affecting the ABD domain. This prevents us from establishing precise genotype-phenotype correlations. Most likely, the specific structural and functional effects of each variant intersect with the genetic background to determine the extent to which tRNA^Phe^ aminoacylation is impacted in each individual and the specific clinical presentation in that individual. For example, of the variants reported here, p.(Arg419His) was previously identified in a patient with pediatric spastic paraplegia and encephalopathy (18). Like this patient, Pt 1 had a non-fatal disease and did not present any epileptic phenotype. Although similar, the two phenotypes were not identical, which may result from the fact that the patient reported by Jou and colleagues was compound heterozygous for this missense variant and a large deletion whereas Pt 1 is compound heterozygous for the p.(Arg419His) variant and a frameshift variant whose expression status is unclear. Pt 3 also expressed the p.(Arg419His) variant but deviated from the other two patients in that he also had epileptic phenotype and white-matter degeneration resulting in a relatively more severe disease. In this case, the p.(Arg419His) variant is compound heterozygous with another missense mutant, p.(His84Pro), that may account for the specific phenotype observed in the patient. Thus, phenotypic differences in these cases may result from different genetic modifiers and/or the presence of different compound heterozygous *FARS2* variants. Keeping in mind the small number of patients, it appears that the p.(Arg419His) variant is associated with long survival with or without epilepsy and white matter abnormalities. In the absence of *in vitro* activity data, we speculate that even though Arg419 is a structurally-important amino acid, its exchange to proline results in a semi-functional enzyme in manner similar to that proposed for p.(Arg419Cys) which resulted in spastic paraplegia and only transient epileptic seizures (13, 19).

Our data further expands the genetic spectrum of *FARS2* variants and raises awareness that some patients may present with early-onset epilepsy and long survival. As many patients with *FARS2* variants are compound heterozygous for two missense variants it would be interesting to integrate data from *in vitro* activity assays of individual and combined mutant mtPheRS to determine to what extent, or if at all, different mutant mtPheRS interact functionally and whether this could add additional information that can help explain the phenotypic presentations observed in different patients.

## Data Availability

Data are available upon request. Variants were submitted to ClinVar

## Acknowledgements

We thank the patients and their families for participating in this study.

